# COVID-19 Outcomes in Saudi Arabia and the UK: A Tale of Two Kingdoms

**DOI:** 10.1101/2020.04.25.20079640

**Authors:** Saleh Komies, Abdulelah M. Aldhahir, Mater Almehmadi, Saeed M. Alghamdi, Ali Alqarni, Tope Oyelade, Jaber S. Alqahtani

## Abstract

**Background:** While the number of COVID-19 cases and deaths around the world is starting to peak, it is essential to point out how different countries manage the outbreak and how different measures and experience resulted in different outcomes. This study aimed to compare the effect of the measures taken by Saudi Arabia and the United Kingdom (UK) governments on the outcome of the COVID-19 pandemic as predicted by a mathematical model.

**Method:** Data on the numbers of cases, deaths and government measures were collected from Saudi’s Ministry of Health and Public Health England. A prediction of the trend of cases, deaths and days to peak was then modelled using the mathematical technique, Exponential Logistic Growth and Susceptible Infectious Recovered (SIR) model. The measures taken by the governments and the predicted outcomes were compared to assess effectiveness.

**Result:** We found over three months that 22 fast and extreme measures had been taken in Saudi Arabia compared to eight slow and late measures in the UK. This resulted in a decline in numbers of current infected cases per day and mortality in Saudi Arabia compared to the UK. Based on the SIR model, the predicted number of COVID-19 cases in Saudi as of 31st of March was 2,064, while the predicted number of cases was 63012 in the UK. In addition, the pandemic is predicted to peak earlier on the 27th of March in Saudi Arabia compared to the 2nd of May 2020 in the UK. The end of transition phases for Saudi and UK according to the model, were predicted to be on 18th of April and 24th of May, respectively. These numbers relate to early and decisive measures adopted by the Saudi government.

**Conclusion:** We show that early extreme measures, informed by science and guided by experience, helped reduce the spread and related deaths from COVID-19 in Saudi. Actions were taken by Saudi under the national slogan “We are all responsible” resulted in the observed reduced number of current and predicted cases and deaths compared to the UK approach “keep calm and carry on”.

## Introduction

SARS-CoV-2 is one species in a family of many coronaviruses that can cause a range of diseases in humans, ranging from the common cold to severe acute respiratory syndrome (1). One of the first pneumonia viruses of COVID-19 was identified in Wuhan city, the capital of Hubei Province in China on 17^th^ of Nov 2019 (2). The first four cases reported were linked to Huanan Seafood Wholesale Market (3). Since then, the disease has spread from Wuhan to other parts of China and worldwide. On the 30^th^ of January 2020, the World Health Organization (WHO) declared the 2019 coronavirus (Covid-19) as a public health emergency of international concern (4), and on the 11^th^ of March 2020, announced that COVID-19 is a global pandemic (4). As of the 24^th^ of April 2020, a total of 2,801,065 laboratory-confirmed cases had been documented globally with a total of 195,218 deaths (5, 6).

Covid-19 is highly communicable, and the average infected persons spread the disease to two or three others resulting in an exponential rate of increase (7). COVID-19 is indeed a massive threat to humanity, and governments across the globe realize that early and decisive actions are more effective in curtailing the disease. However, for most countries, this was not the case. In some countries, early and effective measures have been applied to contain the virus while others delayed or even overlook implementing any decisions. An example of effective early precautionary measures is that taken by the Chinese government which implemented unprecedented non-pharmaceutical interventions to curb the virus including stopping travel from and to Wuhan, the origin of the epidemic as well as 15 other cities in the Hubei province (8, 9). In addition to case isolation, schools and universities were closed, flights and trains were suspended, and roads were closed (8, 9).

Non-pharmaceutical interventions were the first measures that countries across the world implemented. These measures include individual case isolations, the closure of schools and universities, banning of mass gatherings and public events, and most recently, wide-scale social distancing including local and national lockdowns (9). Way before COVID-19 spread to the middle eastern countries; Saudi Arabia implemented measures to prevent the disease from spreading to the country. On the 1^st^ of February, Saudi Arabia stopped flights from and to China before any infected confirmed cases in the kingdom. This was followed by several measures embodied by Saudi’s national slogan *“We are all responsible”*, including but not limited to flights suspension, school closures, case isolation and quarantine for travellers who came back to the country for 14 days in hotels until the decision was made to lockdown the whole country and full travel suspension (6, 10-13). Indeed, the transmission of COVID-19 in Saudi Arabia to date has been low, which reflects the value of applying early precautionary measures.

The United Kingdom government’s response to Covid-19 includes several stages of escalation. The first policy was simple “Keep Calm and Carry On” campaign with mass gathering still allowed without restrictions or distancing. This was clearly stated when the government held its first official press conference on the 12^th^ of March 2020, no recommendations or instructions were announced to stop mass gathering even though the cases of COVID-19 have risen to over 6,000 and mortality above 100 (14, 15). The government’s lack of action continued despite the prime minister’s admission that “many more families will lose loved ones before their time”. On 16^th^ of March, the tone of voice during a second press conference on the same day was different as the prime minister asked the public to avoid pubs, clubs, theatre and other social events as well as non-essential travels (16, 17). He also asked people to work from home were possible, and anyone with persistent cough or fever to self-isolate for seven days (18). Although the government encouraged social distancing, there was no clear directive about mass gatherings. The main decision was only made when a group of researchers at the Imperial College London published a model that predicted that the National Health Service (NHS) may be overwhelmed if no action was taken. This was then immediately followed by a nationwide lockdown, including the closure of schools and universities (19).

Mathematical models have always been used to understand and predict how diseases spread in populations. These models are tools for designing and evaluating the effectiveness of control strategies (20). The mathematical models used in this paper is based on an Exponential Logistic Growth and Susceptible Infectious Recovered (SIR) model, which is a standard compartmental model in epidemiology (21-25). Previously, this model was used to make daily predictions and to estimate the approximate final size of the coronavirus epidemic (22, 26, 27). The model is based on the previous data from the population of interest. Thus, our prediction is based on the data of reported infected and death cases for each country (26, 28, 29). Indeed, the purpose of this study was to analysis the preventative measures implemented by Saudi Arabia and the UK and how these measures affect the reported and predicted cases of COVID-19.

## Method

We reported the trend of COVID-19 (new cases and deaths only) as well as the measures that had been taken by the governments. Data for Saudi Arabia and the UK were sourced from the Ministry of Health Saudi Arabia and Public health of England, respectively (6, 30). Academic data used are available online at the Johns Hopkins University Coronavirus Resource Centre and other sources (1, 12, 30-32).

In terms of data comparison between the two countries, we compare the number of infected and death cases of COVID19 per day for each country. The process of implementing non – pharmaceutical measures and responses actions varies between the two countries. The measures were documented since the beginning of each decision or recommendation by the governments of the UK and KSA.

The expected trends for the two countries are predicted using an algorithm to extrapolate the number of infections and death cases per country (33). The prediction model we used is based on a non-linear coupled differential equation called Susceptible Infectious Recovered (SIR) model, which is a common compartmental model in epidemiology (26, 28). The model is data-driven, thus, the predictions were based on the variation in the data (26, 28, 29). The method is not suitable if the epidemic is still in the early stage. Also, results are not significant if the regression statistic does not meet minimum criteria, say “if R^2^ > 0.8, p-value < 0.05” (26, 28, 29). The algorithm is available online on the MathWorks website and runs on MATLAB program software (29). It was created by Milan Batista to track the spread of Coronavirus (COVID-19) (26, 28). The algorithm models fit data to a logistic model in order to approximate the total number of cases, basic reproduction rate, death rates and expected time scale for the epidemic (26, 28, 29).

### ➢ Prediction Phases

The prediction model we used produces a figure that shows several phases of the epidemic (Fig. 1). For convenience, the evaluation of the epidemic was divided into five regions with four distinct colours and red divider in the middle. Each colour represents different phases of the epidemic.

- White block represents phase one, which follows an exponential growth pattern (slow growth or lag phase): *t* < *t*_*p*_ − 2/*r*
- First orange block represents phase two which describes a fast growth (growth phase or acceleration phase): *t*_*p*_ − 2/*r* < *t* < *t*_*p*_
- Second orange block represents phase three, which is fast to the steady-state growth (negative growth phase or deceleration phase): *t*_*p*_ < *t* < *t*_*p*_ + 2/*r*
- Yellow block represents the phase four, which is the steady-state transition to steady-state phase (slow growth or transition phase): *t*_*p*_ − 2/*r* < *t* < *t*_*p*_
- The green block represents phase five, which is the end phase (plateau stage): *t* > 2*t*_*p*_

The phases are not standard but arbitrarily chosen for convenience (26, 28, 34).

**Figure 1:**
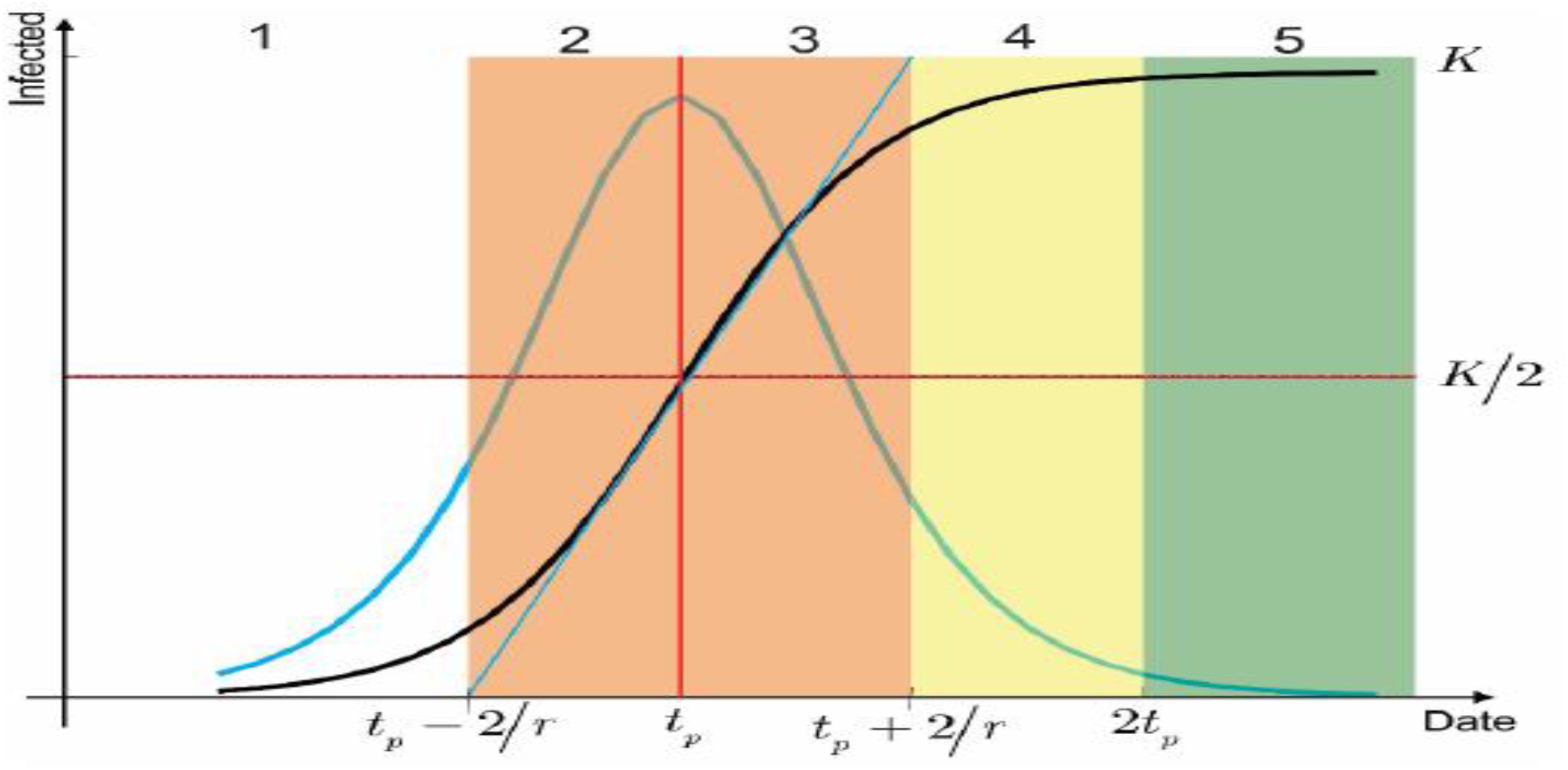
Epidemic Phases (26)

## Results

### ➢ Current infected cases in Saudi Arabia and the UK

The government of the UK implemented the first measure after the 18^th^ day of the first confirmed case of COVID-19, whereas Saudi Arabia’s first measure was 28 days before the first confirmed case of COVID-19. The number of confirmed cases in line with the government’s measures in response to the pandemic over three months (January through March 2020) in Saudi Arabia and the UK is shown in Figure 2. In general, Saudi Arabia had a lower number of infected cases per day compared to the UK. As shown, the Saudi government announced 22 consecutive measures to prevent and contain the spread of the pandemic. Compared to Saudi Arabia, the UK implemented eight measures over the same time. Over three months, the total numbers of confirmed cases in KSA and UK were 1,720 versus 29,461, respectively. A series of declines in the new confirmed cases in Saudi Arabia after 21 days from the date of the first case is shown in Figure 2. In contrast, the number of new cases in the UK was still rising, with no further measures announced by the UK government. The UK takes longer time than KSA to reach the peak of confirmed new cases of COVID-19, as shown in Figure 2. Conversely, government measures played a vital role to minimize the number of new cases every day. The relationship between the number of measures and the number of new cases in both countries could not be tested due to limited data.

**Figure 2:**
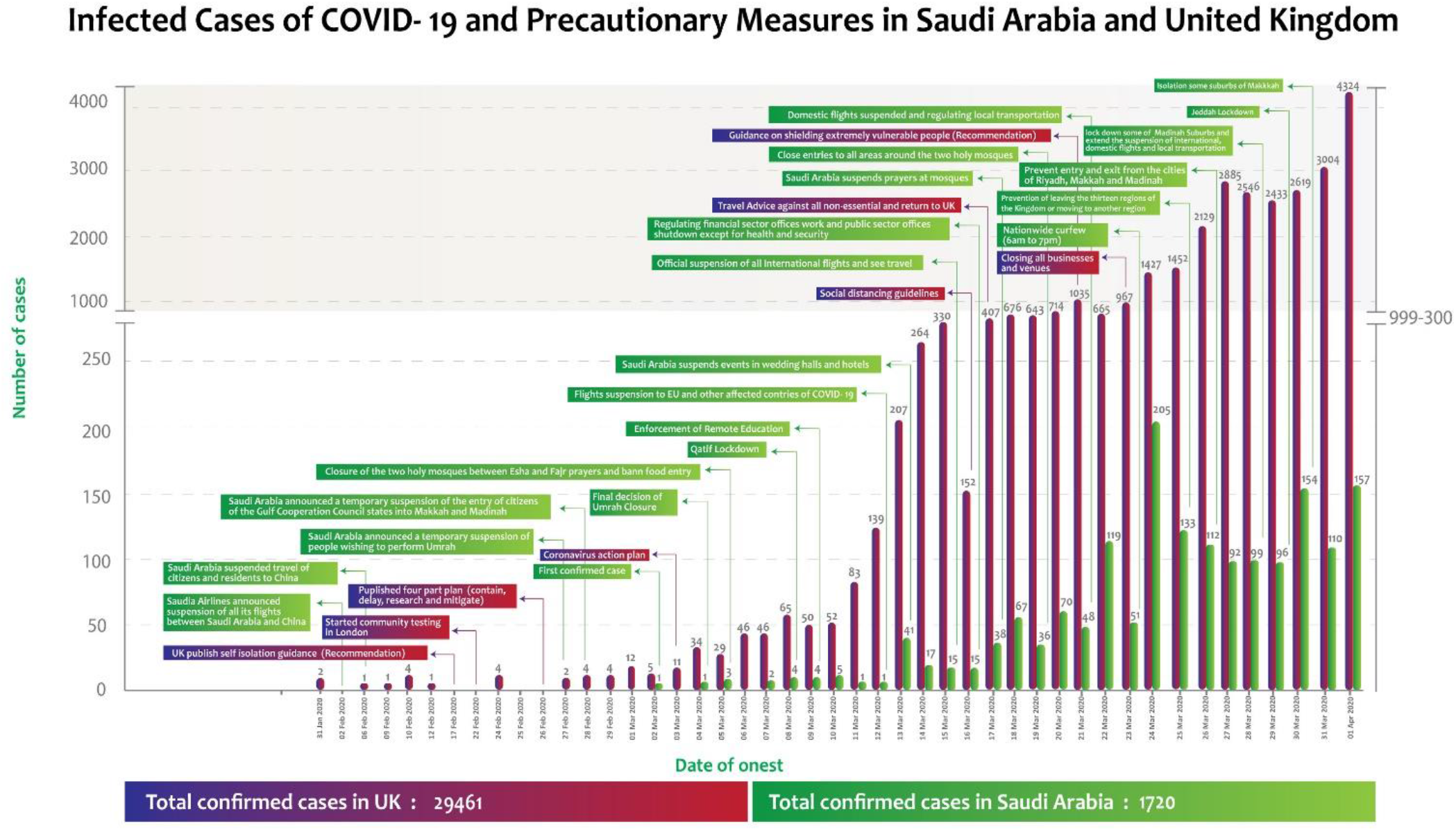
Confirmed Cases of COVID-19 and Measures in Saudi Arabia and the United Kingdom.

### ➢ Current death cases in Saudi Arabia and the UK

Figure 3 shows a summary of the number of deaths in conjunction with the governmental measures due to COVID-19 over three months in KSA and the UK. The total number of deaths in KSA was lower than the number of deaths in the UK (16 versus 2,352). The date of onset started late in March in KSA, compared to the UK. Also, there was a consistent pattern in the number of deaths in KSA compared to the UK. The onset of death due to COVID-19 in the UK was earlier in March. UK death cases had a dramatic increase over three months compare to KSA. In total, KSA announced 16 measures to be taken in order to control the deaths due to COVID-19. In total, the UK announced only four measures to be taken in order to control the deaths due to COVID-19. There was limited data to test the relationship between the number of measures and the number of deaths in both countries.

**Figure 3.**
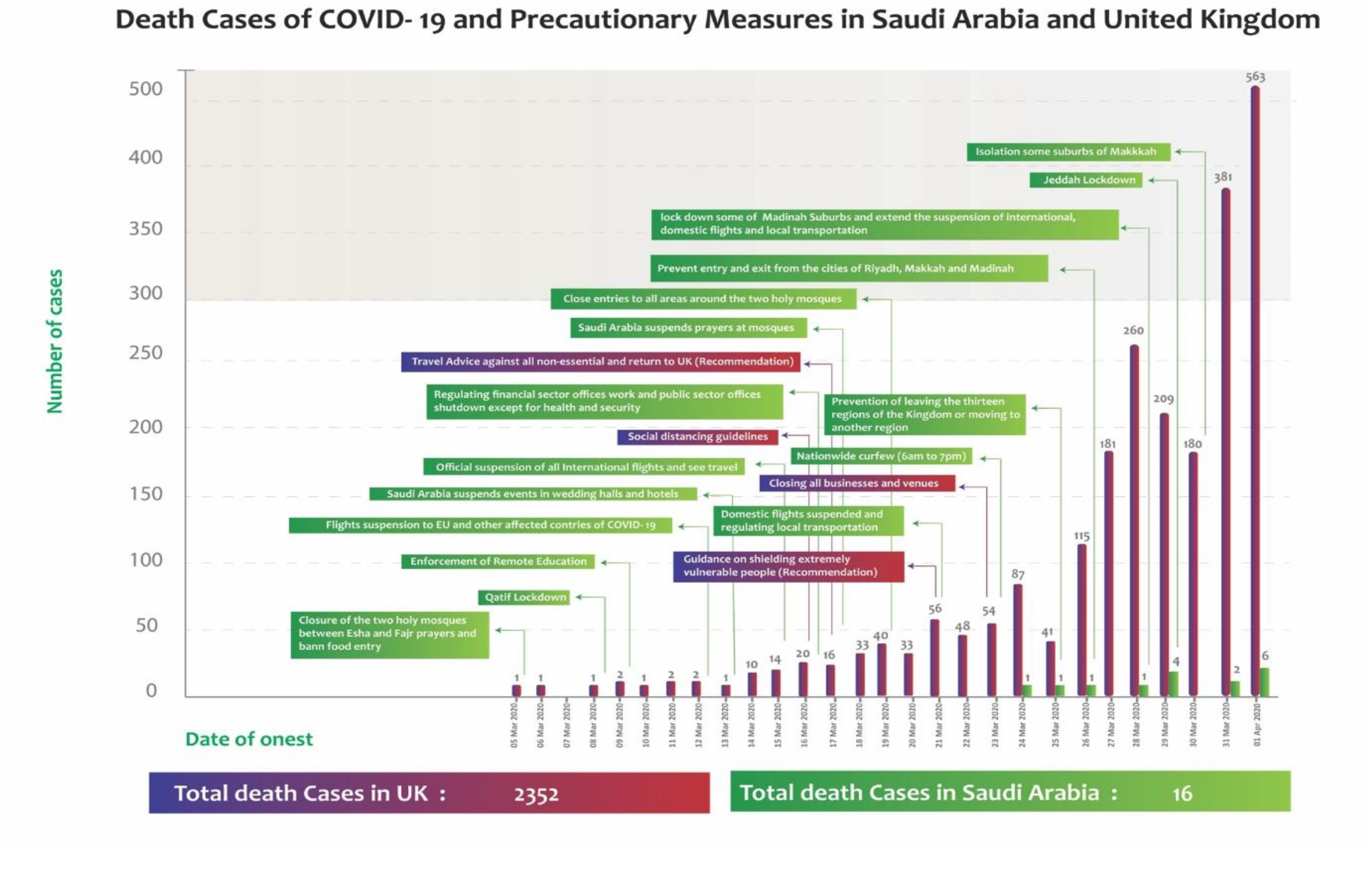
Death Cases of COVID-19 and Measures in Saudi Arabia and the UK.

### ➢ Predicted infected and death cases in Saudi Arabia

The model was tested on the 1^st^ of April 2020, 31 days after Saudi Arabia reported the first case. The predicted trend (infections per day) of COVID-19 in Saudi Arabia can be seen in Figure 4. When we run the model, the actual number of cases and deaths was 1,720 and 16 respectively. After the first case, the infection rate and the number of cases started to increase albeit slowly, taking 17 days to peak (n=205). According to Figure 4, the trend of new cases is consistent with the control measures put in place in Saudi and the rise in number was not as fast as in the UK. Based on the logistic model, the approximate final size by the 31^st^ of March, of coronaviruses cases in Saudi Arabia would be 2,064. The peak of the epidemic was expected on the 27^th^ of March 2020. The model also predicted that the end of the transition phase will be around the 18^th^ of April 2020 (the region between yellow and green). (Figure 4)

**Figure 4:**
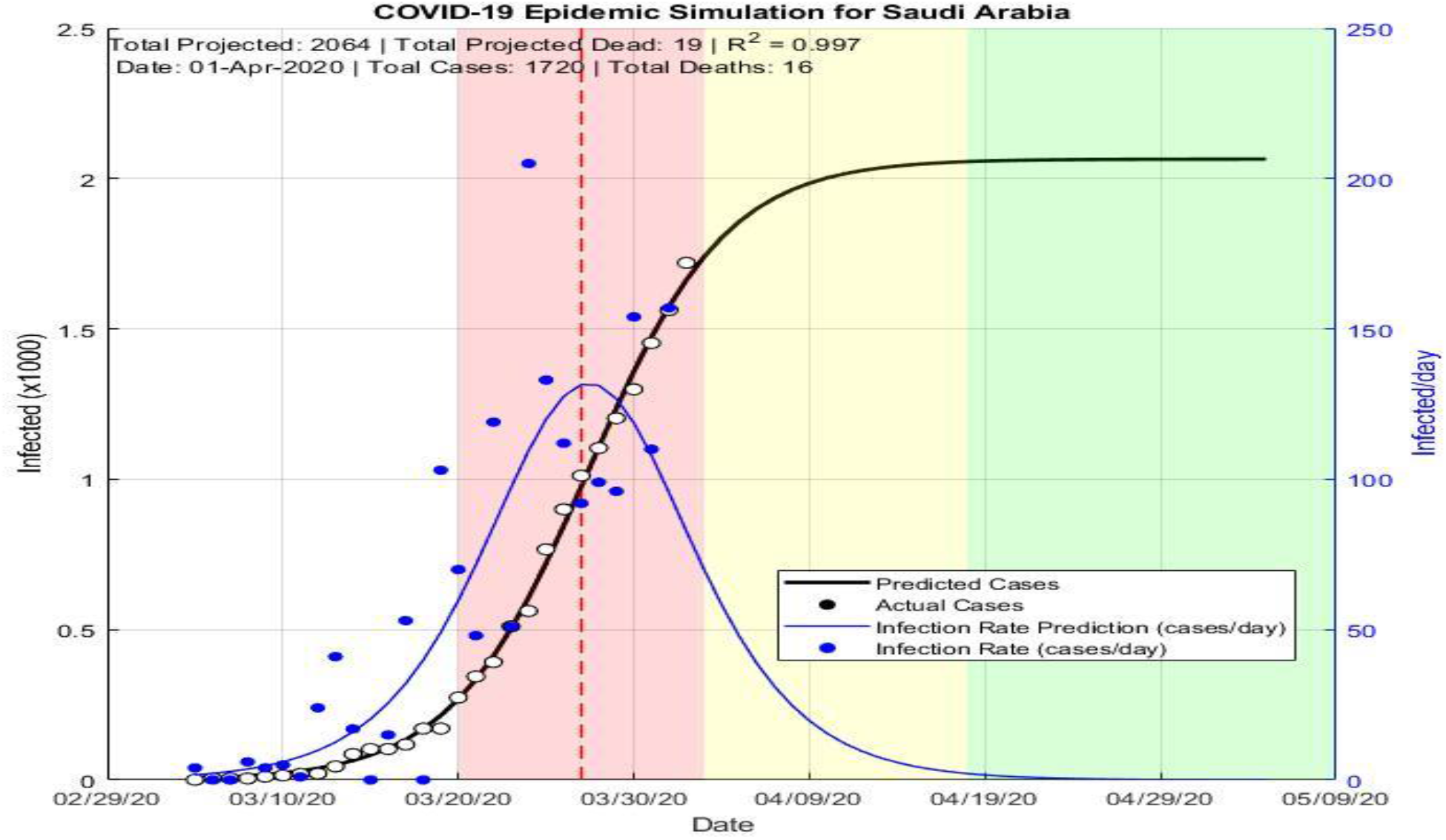
Predicted evaluation of coronavirus epidemic in Saudi Arabia.

In addition, the logistic regression model and estimated coefficients for Saudi Arabia is shown in Table 1:

**Table 1:** Estimated logistic parameters for Saudi Arabia data is up to first of April 2020. Estimated Coefficients parameters for Saudi Arabia:

|  | Estimate | SE | tStat | pValue |
| --- | --- | --- | --- | --- |
| K | 2064.7 | 74.731 | 27.629 | 8.4551e-21 |
| r | 0.25557 | 0.0098014 | 26.075 | 3.6205e-20 |
| A | 309.12 | 45.903 | 6.7342 | 3.8084e-07 |

Number of observations: 29, Error degrees of freedom: 26
Root Mean Squared Error: 28.6
R-Squared: 0.998, Adjusted R-Squared 0.997
F-statistic vs. zero model: 6.24e+03, p-value = 2.94e-37

**Table 2:**
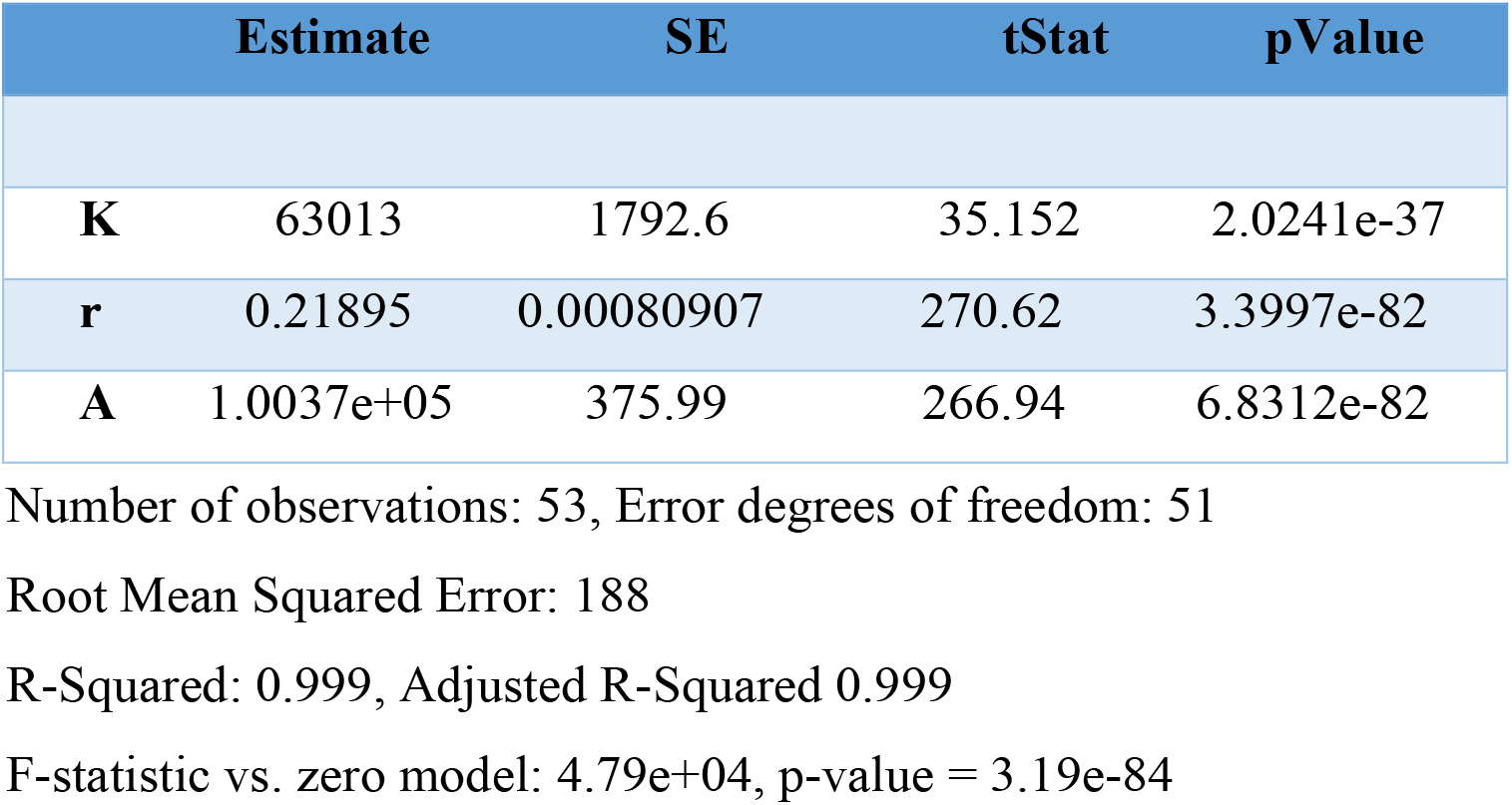
Estimated logistic parameters for United Kingdom data is up to first of April 2020 Estimated Coefficients parameters for the UK

**Table 3:** Precautionary Measures and Government’s Response of Saudi Arabia

| **Date** | **Decision** |
| --- | --- |
| 02/02/2020 | Saudi Arabian Airlines announced the suspension of all its flights between Riyadh and Jeddah to and from Guangzhou |
| 06/02/2020 | Saudi Arabia suspended travel of citizens and residents to China |
| 27/02/2020 | Saudi Arabia announced a temporary suspension of entry for individuals wishing to perform Umrah in Mecca or visit the Prophet's Mosque in Medina, as well as tourists. The order was also expanded to include visitors traveling from countries where the COVID-19 is a threat. |
| 28/02/2020 | The Minister of Foreign Affairs of the Kingdom of Saudi Arabia announced a temporary suspension entry of citizens of the Gulf Cooperation Council states into Makkah and Madinah. Citizens of the countries of the Gulf Cooperation Council who have been in the Kingdom of Saudi Arabia for more than 14 consecutive days and who have not had any symptoms of the Covid-19 will be excluded from the decision |
| 02/03/2020 | Closure of both mosques (Mecca and Medina) between the evening Esha prayers and morning Fair prayers, and a ban on food entering the two complexes. |
| 08/02/2020 | Temporarily suspending entry and exit to Qatif |
| 09/03/2020 | The Ministry of Education announced that it will temporarily suspend studies in all regions and governorates of the Kingdom. The decision includes schools, public, private and university education institutions and the public and private technical and vocational training institution. |
| 12/03/2020 | Suspending travel of citizens and residents and suspending flights to the European Union, Switzerland, India, Pakistan, Sri Lanka, the Philippines, Sudan, Ethiopia, South Sudan, Eritrea, Kenya, Djibouti and Somalia, with the exception of health practitioners working in the Kingdom from the countries of the Philippines and India. The movement of passengers was also suspended through all land ports with Jordan except for commercial traffic, freight and humanitarian and exceptional cases. |
| 13/03/2020 | Saudi Arabia suspends events in wedding halls, rest houses, and hotels |
| 15/03/2020 | Saudi Arabia suspend international flights for two weeks and The Ministry of Sports announced the suspension of all sports activities in the Kingdom of Saudi Arabia in all tournaments and competitions, as well as the closure of private sports halls and centres, and |
| 16/03/2020 | Suspend attendance at workplaces in all government agencies for 16 days, except for health, security, and military sectors. |
| 17/03/2020 | Saudi Arabia suspends prayers at mosques |
| 19/03/2020 | Suspending all Vehicles from entering the parking lot of the Prophet's Mosque in Medina temporarily and Suspending the presence and prayer in the grounds of the Two Holy Mosques, especially on Friday, and suspending the attendance of worshipers for prayers in the Prophet's Mosque |
| 21/03/2020 | Saudi Arabia suspended on Friday all domestic flights, buses, taxis and trains for 14 days and Shops are closed from eight in the evening until six in the morning, except for food stores, pharmacies and gas stations |
| 23/03/2020 | A daily curfew, from 7pm to 6am, for 21 days |
| 25/03/2020 | Prevention of leaving the thirteen regions of the Kingdom or moving to another region |
| 26/03/2020 | Prevent entry and exit from the cities of Riyadh, Makkah and Madinah, according to the limits set by the concerned authority. |
| 28/03/2020 | Preventing entry or exit from 6 districts in Madinah for a period of 24 hours until ensuring that there are no cases that require dealing with or the end of the medically recommended isolation period |
| 29/03/2020 | Suspending entry and exit from Jeddah Governorate, and submitting a curfew date to start from 03:00pm |
| 30/03/2020 | Isolation of neighbourhoods: Ajyad, Al-Masafi, Al-Masfalah, Al-Hujun, Al-Nukseh and Hosh Al-Bakr, and preventing entry to or exit from it and extending curfews to 24 hours. |

**Table 4:**
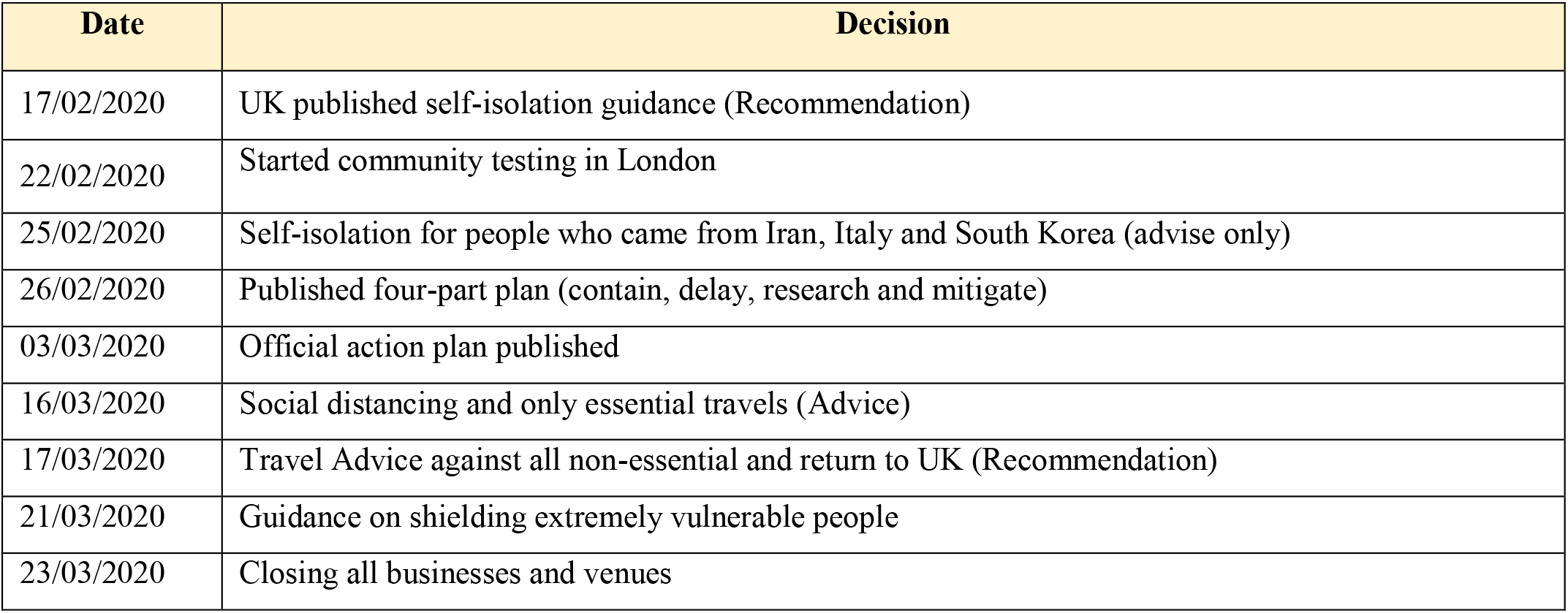
Precautionary Measures and Government’s Response of the UK

| Date | Decision |
| --- | --- |
| 17/02/2020 | UK published self-isolation guidance (Recommendation) |
| 22/02/2020 | Started community testing in London |
| 25/02/2020 | Self-isolation for people who came from Iran, Italy and South Korea (advise only) |
| 26/02/2020 | Published four-part plan (contain, delay, research and mitigate) |
| 03/03/2020 | Official action plan published |
| 16/03/2020 | Social distancing and only essential travels (Advice) |
| 17/03/2020 | Travel Advice against all non-essential and return to UK (Recommendation) |
| 21/03/2020 | Guidance on shielding extremely vulnerable people |
| 23/03/2020 | Closing all businesses and venues |

### ➢ Predicted infected and death cases in the UK

The model was tested on the 1^st^ of April 2020, which is the 62^nd^ day of the epidemic. The predicted numbers of COVID-19 cases and death in the United Kingdom is presented in Figure 5. At that time, we run the model, and the number of cases and deaths were 2,9461 and 2,350 respectively. The first case of COVID-19 in the UK was reported in late January 2020 after which the number of new cases surged dramatically. Accordingly, it took the UK two months to reach the peak number of cases per day (n=4400), with the number of daily deaths taking a relatively shorter time to jump from hundreds to thousands. Based on the logistic model, the estimated total number of coronaviruses cases in the UK would be around 63,012. Also, the peak of the epidemic was expected on the 2^nd^ of April 2020 and estimated end of transition phase on the 24^th^ of May 2020 (the region between yellow and green).

**Figure 5.**
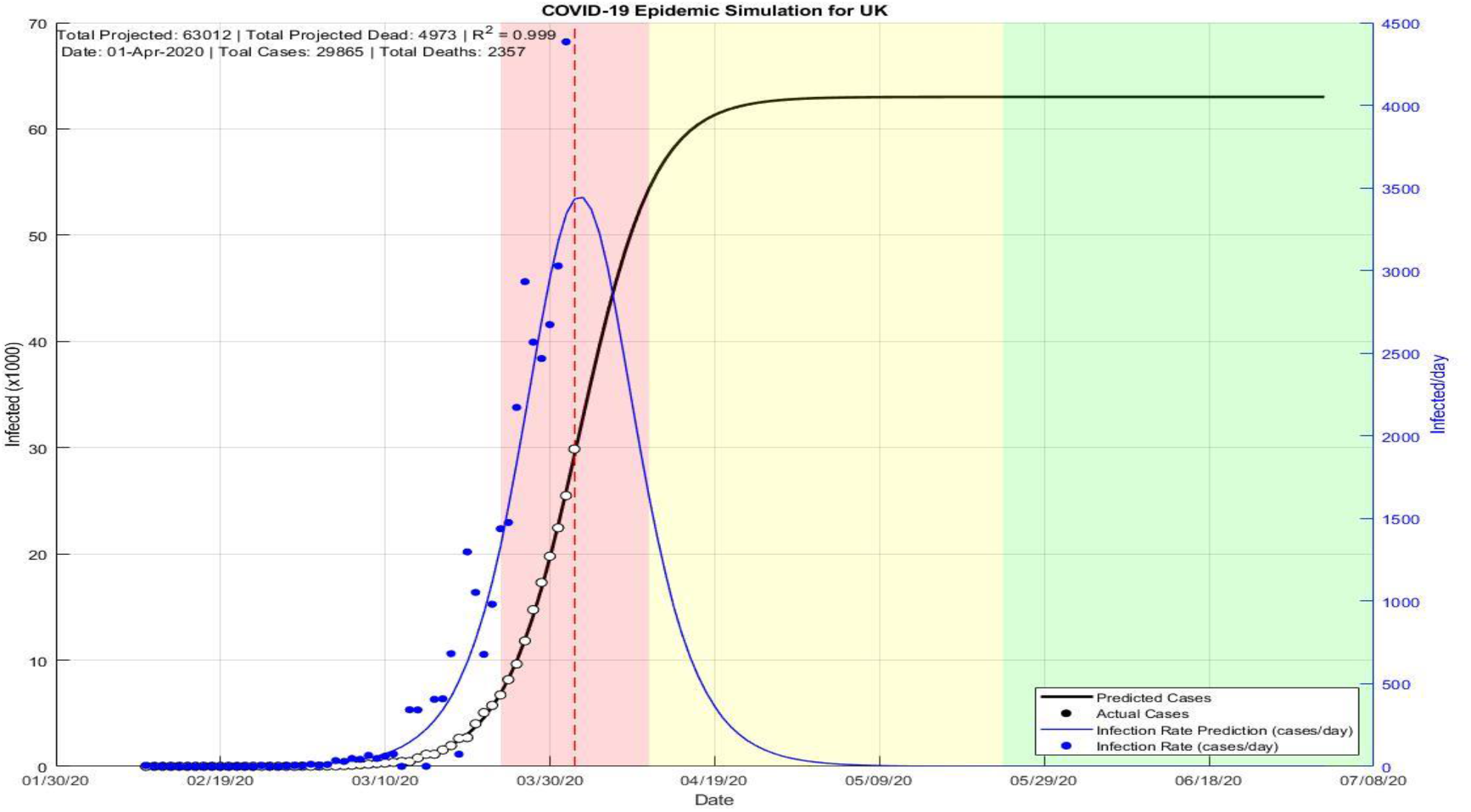
Predicted evaluation of coronavirus epidemic in United Kingdom.

#### Precautionary Measures of Saudi Arabia

The table below presents all the precautionary measures in details per date for Saudi Arabia (6, 12, 13).

### ➢ Precautionary Measures in the UK

The table below presents all the precautionary measures and recommendation versus date for the UK (14, 16, 30).

## Discussion

For the first time, we compare how the Covid-19 outbreak was tackled in Saudi Arabia and the UK. This included current infected cases, death cases, predicted outbreak and government’s response in both countries. We found over three months that 22 quick and decisive measures were taken in Saudi Arabia compared to eight slow and lacklustre measures in the UK. This resulted in a decline in numbers of infected cases per day, mortality, and predicted new cases with their associated deaths in Saudi Arabia compared to the UK.

WHO declared the COVID-19 outbreak a “Public Health Emergency of International Concern” on 30 January 2020 (35). Early before the first case was reported on the 2^nd^ March 2020, Saudi Arabia demonstrated high preparedness by undertaking exceptional precautionary measures. The Saudi government promoted a policy of *“We are all responsible”* to the public, encouraging and enforcing compliance with the guidelines provided by the authorities to counter the spread of COVID-19 (6, 12, 13).

On the part of Saudi Arabia, pre-emptive measures were put in place, which prioritize the safety of the citizen with a full understanding of the social, religious and financial implications. Two days after the declaration by WHO of a global emergency, Saudi Arabia suspended all travels to and from Guangzhou, China. This was swiftly followed by an array of measures including the suspension of travels for citizens and residents of China, temporary suspension of the holy hajj and Umrah pilgrimage, and closure of borders shared with other Gulf countries. It is important to note that these measures were implemented before the first case was reported on the 2^nd^ of March 2020. Conversely, between the time of declaration of COVID-19 as a global emergency by WHO (30 of January) and after the first UK cases (31 of January) no action was taken in the UK. The UK Government did not take any precautionary actions until the 17^th^ of February 2020, when a set of recommendations on self-isolation was published (14, 16, 30). All the while, flight between China and the UK were operating at full capacity with pubs and business venues still open. This left a hiatus of over two weeks in which time life continued as usual with congested transport systems continuing to run and people still gathering in masses.

The first case was reported in the eastern province of Saudi Arabia on the 2^nd^ of March. This triggered another series of concerted actions by the Saudi’s government. On the same day, all mosques in Mecca and Medina were closed followed a week later by the lockdown of the whole eastern province. Between the 9^th^ and 12^th^ of March, studies in schools and universities were suspended and flights to and from 13 nations including the European Union suspended for all residents and citizens. The land ports between Saudi and Jordan was closed. From the 13^th^ to 21^st^ of March, mass gatherings in the form of weddings, sporting events, workplaces, mosque prayers (including on the Prophet’s mosque), and public transport were suspended. These were followed up on the 23^rd^ of March by a combination of six measures including curfews and regional isolation. All in all, Saudi followed and exceeded the WHO recommendations (6, 12, 13). As of 24^th^ of April, currently confirmed cases in Saudi Arabia stands at 15,102 with 127 deaths, which indicates less transmission and consequently less mortality.

On the 31^st^ of January 2020, the UK government reported its first cases when two Chinese nationals from the same family test positive for the SARS-CoV-2. Early at this stage, the UK government initially adopted the WHO recommendation of contact tracing and isolation of people with proven exposure. However, this was abandoned, and mass gathering was still going on, including sporting events until the 20^th^ of March when the UK government officially announced a national lockdown (36). By this time, the total number of infection had already gone above 6,000 with 48 COVID-19-related deaths (37). The delayed actions by the UK government against a background of a population with a higher proportion of older and high-risk people makes the UK susceptible to higher infection and death from COVID-19 (38, 39). This difference in approach may explain the difference in the total number of actual and predicted cases and death presented in this study. This may also explain the difference in the predicted days for the infection to peak according to the logistic model (Saudi = 17 days, 220 cases; UK = 2 months, 4400 cases). As of 24^th^ of April, currently confirmed cases in the UK were 143,464 with 19,506 deaths, which shows high transmission and death rate compared to Saudi (32). However, the high death rate reported in the UK may also be linked to limited access to respiratory support as part of COVID-19 management, the severity of the respiratory disease, and critical care capacity in each hospital or region (40-43).

Taken together, this shows a lack of preparedness by the UK government in preventing the spread and burden of COVID-19, and this is evident by the total number of infected cases and deaths compared to Saudi. *“Keep Calm and Carry On”* was the public face of the UK government’s response to Covid-19 outbreak (44). Nonetheless, this claim was based on the behavioural sciences viewpoint that “fatigue” with stringent COVID-19 control measures would be developed if they were activated too early (44). Still decisions of this magnitude should have been made based on scientific evidence especially as the UK has the fourth-lowest number of hospital beds per 1000 population among the G20 nations (45). The combined actions by the Saudi Arabia’s government showed a high level of preparedness coupled with sound advice guided by science. These decisions were based and informed by extensive experience in the control of previous pandemic (MERS) (46, 47). The aim was to reduce both the economic and human cost by taking early decisive and definitive actions understanding fully that early sacrifices will reduce the burden to the national health infrastructures and pressure on healthcare workers.

It is worth mentioning that the logistic model has some drawback, especially when approaching the final stage of the epidemic. More specifically, when the real number of cases is slightly larger than the number predicted by our model or it exceeded the predicted end-stage systematically. Another drawback is that the phase of the epidemic is not described by the SIR model; thus the model cannot be extrapolated unless the same arbitrary format is used (26, 28, 34). Generally, the prediction model for new cases for both Saudi Arabia and the UK was not robust enough as it only explained 25% in Saudi Arabia, and 21% in the UK. Both models were generated for 31 days from the beginning to 1^st^ of April 2020. The model did not account for confounders such as age and gender fractions, governmental measures, and cities locked down. However, it assumes a uniform mixing of the population, where recovery is equally likely among the infected population (48). The pattern of recovery and mortality rates in Saudi Arabia as well as in UK were not estimated due to incomplete data. Importantly, comparing infected cases in Saudi Arabia to the UK demonstrated a dramatic increase in UK. Similarly, the number of deaths in Saudi Arabia are less than the number of deaths in the UK. That may be due to the UK’s higher population and population density compared to Saudi. However, the death per 1000 individual in Saudi is still significantly lower when controlled for population size (UK = 4.6×10^−4^; Saudi = 3.5×10^−2^). Thus, this difference might be related to the different governmental approaches taken at the time.

These findings contribute in several ways to our understanding of COVID-19 response in both kingdoms and provide a groundwork for future research. To our knowledge, this work is the first to compare the effects of two countries’ measures on the outcome of a global pandemic with a specific focus on how decisive and early actions helped reduce the number of cases and deaths recorded. Although the results of this study were based on previously reported data from both countries, when applied to current data, the model still showed positive results favouring early measures as was taken by Saudi Arabia. Because this model relies on early reported cases, it might serve as a resource for decision making regarding future pandemics. Also, as telemedicine have been effective in the delivery of care to individuals in their home (49, 50), future studies should investigate the effectiveness of using telemedicine for Covid-19 patients in both countries and how this affects the pandemic outcomes.

## Conclusion

We show that early decisive measures informed by science and guided by experience can help reduce the spread and related death from infectious diseases. Actions were taken by Saudi under the national slogan *“We are all responsible”* resulted in the observed reduced number of cases and deaths compared to the UK approach *“keep calm and carry on”*. This strategy adopted by the Saudi authorities may serve as a blueprint for the management of potential future pandemics.

## Data Availability

NA

## Acknowledgement

- The authors would like to thank Dr. Metib Alghamdi from King Khaled University (Saudi Arabia) and Joshua McGee from The University of Massachusetts Amherst (United State) for their helpful advice on various technical issues that was encountered.

## References

1. Organization WH. Coronavirus disease (COVID-19) outbreak 2020 [Available from: https://www.who.int/csr/disease/coronavirus_infections/ar/.

2. Davidson H. First Covid-19 case happened in November, China government records show - report 2020 [updated 13/03/2020. Available from: https://www.theguardian.com/world/2020/mar/13/first-covid-19-case-happened-in-november-china-government-records-show-report.

3. Li Q, Guan X, Wu P, Wang X, Zhou L, Tong Y, et al. Early Transmission Dynamics in Wuhan, China, of Novel Coronavirus–Infected Pneumonia. New England Journal of Medicine. 2020;382(13):1199–207.

4. Organization WH. Coronavirus disease (COVID-19) 2020 [Available from: https://www.who.int/emergencies/diseases/novel-coronavirus-2019/events-as-they-happen

5. Orgnization WH. Coronavirus disease 2019 (COVID-19) Situation Report – 72: WHO 2020 [updated 01/04/2020. Available from: https://www.who.int/docs/default-source/coronaviruse/situation-reports/20200401-sitrep-72-covid-19.pdf?sfvrsn=3dd8971b_2.

6. Arabia MoHoS. Media Center 2020 [updated 01/04/2020. Available from: https://www.moh.gov.sa/en/Ministry/MediaCenter/Pages/default.aspx.

7. Gates B. Responding to Covid-19 - A Once-in-a-Century Pandemic? The New England journal of medicine. 2020.

8. Cyranoski D. What China’s coronavirus response can teach the rest of the world. Nature. 2020:Nature 579, 479–80 (2020).

9. Seth Flaxman SM AGea. Estimating the number of infections and the impact of nonpharmaceutical interventions on COVID-19 in 11 European countries. Imperial College London. 2020.

10. Reutors. Saudi Arabia temporarily suspends entry of GCC citizens to Mecca and Medina: foreign ministry 2020 [updated 28/02/2020. Available from: https://web.archive.org/web/20200229004835/https://www.reuters.com/article/us-health-china-saudi-idUSKCN20M31T.

11. News A. Saudi Arabia announces first case of coronavirus 2020 [updated 03/31/2020. Available from: https://www.arabnews.com/node/1635781/saudi-arabia.

12. Arabia MoIoS. Security spokesman’s statements. Saudi Arabia 2020.

13. Agency PS. Genral Transmissions Press Saudi Agency 2020 [updated 31/03/2020. Available from: https://www.spa.gov.sa/newsheadlines.php?lang=en#page=1.

14. GOV.UK. COVID-19 - guidance for households with possible coronavirus infection 2020 [updated 12 March 2020. Available from: https://www.gov.uk/government/publications/covid-19-stay-at-home-guidance.

15. government U. COVID-19: guidance for households with possible coronavirus infection. 2020

16. GOV.UK. COVID-19 guidance for mass gatherings 2020 [updated 16 March 2020. Available from: https://www.gov.uk/guidance/covid-19-guidance-for-mass-gatherings.

17. government U. COVID-19 guidance for mass gatherings. 2020.

18. Hunter DJ. Covid-19 and the Stiff Upper Lip — The Pandemic Response in the United Kingdom. New England Journal of Medicine. 2020.

19. Neil M Ferguson DL GN-G, Natsuko Imai, Kylie Ainslie, Marc Baguelin, et al. Impact of non-pharmaceutical interventions (NPIs) to reduce COVID19 mortality and healthcare demand. Imperial College London. 2020.

20. Chowell G, Sattenspiel L, Bansal S, Viboud C. Mathematical models to characterize early epidemic growth: A review. Phys Life Rev. 2016;18:66–97.

21. University R. Environmental Limits to Population Growth 2017. Available from: https://cnx.org/contents/GFy_h8cu@10.12:eeuvGg4a@4/Environmental-Limits-to-Population-Growth.

22. Keeling MJ, Danon L. Mathematical modelling of infectious diseases. British Medical Bulletin. 2009;92(1):33–42.

23. Zurich E. SIR models of epidemics 2020 [Available from: https://tb.ethz.ch/education/learningmaterials/modelingcourse/level-1-modules/SIR.html.

24. Biology O. Environmental Limits to Population Growth 2020 [updated Mar 21, 2018. Available from: https://cnx.org/contents/GFy_h8cu@10.12:eeuvGg4a@4/Environmental-Limits-to-Population-Growth.

25. Science EZ-DoES. Theortical Biology - SIR models of epidemics 2020 [Available from: https://tb.ethz.ch/education/learningmaterials/modelingcourse/level-1-modules/SIR.html.

26. Batista M. Estimation of the final size of coronavirus epidemic by the logistic model. 2020. medRxiv. https://doi.org/10.1101/2020.02.16.20023606

27. batista m. Estimation of the final size of the second phase of the coronavirus epidemic by the logistic model. medRxiv. 2020:2020.03.11.20024901.

28. Batista M. Estimation of the final size of the COVID-19 epidemic. medRxiv. 2020:2020.02.16.20023606.

29. McGee J. fitVirusCV19v3 (COVID-19 SIR Model). JosMATLAB Central File Exchange. 2020.

30. England PH. Number of coronavirus (COVID-19) cases and risk in the UK 2020 [updated 24/01/2020. Available from: https://www.gov.uk/government/organisations/public-health-england.

31. Center JHCR. Coronavirus COVID-19 Global Cases by the Center for Systems Science and Engineering (CSSE) at Johns Hopkins University (JHU) 2020 [updated 01/04/2020. Available from: https://coronavirus.jhu.edu/map.html.

32. Worldometer. COVID-19 Coronavirus Pandemic 2020 [updated 07/04/2020. Available from: https://www.worldometers.info/coronavirus/.

33. Langel W. Extrapolation of Infection Data for the CoVid-19 Virus and Estimate of the Pandemic Time Scale. medRxiv. 2020:2020.03.26.20044081.

34. Batista M. Forecasting of final COVID-19 epidemic size. 2020.

35. WHO. Coronavirus disease (COVID-19) technical guidance: laboratory testing for 2019-nCoV in humans (updated 2 Mar 2020). [Available from: https://www.who.int/emergencies/diseases/novel-coronavirus-2019/technical-guidance/laboratory-guidance.

36. Pollock AM, Roderick P, Cheng K, Pankhania B. Covid-19: why is the UK government ignoring WHO’s advice? : British Medical Journal Publishing Group; 2020.

37. Number of coronavirus (COVID-19) deaths in Europe since February 2020, by country Published by Conor Stewart. [Available from: https://www.statista.com/statistics/1102288/coronavirus-deaths-development-europe/.

38. Alqahtani JS, Oyelade T, Aldhahir AM, Alghamdi SM, Almehmadi M, Alqahtani AS, et al. Prevalence, Severity and Mortality associated with COPD and Smoking in patients with COVID-19: A Rapid Systematic Review and Meta-Analysis. medRxiv. 2020: https://doi.org/10.1101/2020.03.25.20043745

39. Yang J, Zheng Y, Gou X, Pu K, Chen Z, Guo Q, et al. Prevalence of comorbidities in the novel Wuhan coronavirus (COVID-19) infection: a systematic review and meta-analysis. International Journal of Infectious Diseases. 2020;12:12.

40. Paterlini M. On the front lines of coronavirus: the Italian response to covid-19. Bmj. 2020;368:m1065.

41. Rosenbaum L. Facing Covid-19 in Italy — Ethics, Logistics, and Therapeutics on the Epidemic’s Front Line. New England Journal of Medicine. 2020.

42. Sreedharan J, Alqahtani J. Driving pressure: Clinical applications and implications in the intensive care units. Indian Journal of Respiratory Care. 2018;7(2):62–6.

43. Oyelade, T.; Alqahtani, J., Canciani, G. Prognosis of COVID-19 in Patients with Liver and Kidney Diseases: An Early Systematic Review and Meta-Analysis. Preprints 2020, 2020040464 (doi: 10.20944/preprints202004.0464.v1).

44. Hunter DJ. Covid-19 and the Stiff Upper Lip—The Pandemic Response in the United Kingdom. New England Journal of Medicine. 2020.

45. Organisation for Economic Co-operation and Development. OECD data: hospital beds. 2020. [Available from: https://data.oecd.org/healtheqt/hospital-beds.htm.

46. Barry M, Ghonem L, Alsharidi A, Alanazi A, Alotaibi NH, Al-Shahrani FS, et al. Coronavirus Disease 2019 (COVID-19) Pandemic in the Kingdom of Saudi Arabia: Mitigation Measures and Hospitals Preparedness.

47. Algaissi, A., Alharbi, N., Hassanain, M., Hashem, A. Preparedness and Response to COVID-19 in Saudi Arabia: Lessons Learned from MERS-CoV. Preprints 2020, 2020040018 (doi: 10.20944/preprints202004.0018.v1)

48. Batista M. Estimation of coronavirus COVID-19 epidemic evaluation by the SIR model 2020 [Available from: https://uk.mathworks.com/matlabcentral/fileexchange/74658-fitviruscovid19?s_tid=prof_contriblnk.

49. Smith AC, Thomas E, Snoswell CL, Haydon H, Mehrotra A, Clemensen J, et al. Telehealth for global emergencies: Implications for coronavirus disease 2019 (COVID-19). Journal of telemedicine and telecare. 2020:1357633x20916567.

50. Alrajeh AM, Aldabayan YS, Aldhair AM, Pickett E, Quaderi SA, Alqahtani JS, et al. Global use, utility, and methods of tele-health in COPD: a health care provider survey. Int J Chron Obstruct Pulmon Dis. 2019;14:1713–9.

